# Diagnostic and Prognostic Value of Global Longitudinal Strain in Heart Failure with Preserved Ejection Fraction: A Systematic Review and Meta-Analysis

**DOI:** 10.1101/2023.03.13.23287207

**Authors:** Yujiao Shi, Wang Qing, Chunqiu Liu, Xiong Shuang, Chenguang Yang, Wenbo Qiao, Guoju Dong, Jiangang Liu

## Abstract

**Objective:** Heart failure with preserved ejection fraction (HFpEF), an increasing public health concern, is increasing in prevalence and is associated with an elevated risk of hospitalization and mortality. Currently, data on the clinical application value of left ventricular global longitudinal strain (LV GLS) in HFpEF are contradictory. Therefore, we performed the following meta-analysis to appraise the diagnostic and prognostic value of LV GLS in HFpEF.

**Methods:** PubMed, Medline, Scopus, and Web of Science were retrieved exhaustively from their inception until December 20, 2022, to obtain literature assessing the diagnostic and prognostic value of LV GLS in HFpEF.

**Results:** Forty-one studies (including 14,543 patients) published from 2008 to 2022 were included. The results of the meta-analysis were as follows: First, the LV GLS values in HFpEF patients were significantly lower than in healthy individuals (SMD:1.21; 95% CI (0.94, 1.47), p<0.00001, I^2^=85%; P<0.00001), but substantially higher than in HErEF patients (SMD: -2.03; 95% CI (−2.23, -1.72), p<0.00001, I^2^=92%; P<0.00001). Second, the pooled diagnostic parameters of LV GLS for HFpEF were as follows: sensitivity, 0.77 (95% CI: 0.71–0.82); specificity, 0.66 (95% CI: 0.58–0.74); DOR, 7.53 (95% CI: 3.19–17.74); AUC for the SROC, 0.81 (95% CI: 0.79–0.87). Finally, the low LV GLS values were correlated with a higher risk of all-cause death (HR: 1.12; 95% CI: 1.01-1.25; p=0.000, I^2^=84%; P = 0.031) in HFpEF patients.

**Conclusions:** LV GLS is impaired in HFpEF patients despite a normal left ventricular ejection fraction, indicating the existence of mild LV contractile dysfunction. Moreover, LV GLS might be an auxiliary indicator for diagnosing HFpEF and predicting all-cause death in HFpEF patients.

## 1 Introduction

Heart failure (HF) with preserved ejection fraction (HFpEF), a complex and heterogeneous ailment, is a multi-organ disorder caused by an aging populace and comorbidities such as obesity, hypertension, diabetes, and chronic renal insufficiency[1]. HFpEF, comprising approximately half of the population of HF, is increasing in prevalence and is associated with an elevated risk of hospitalization and mortality[2,3]. Nevertheless, the clinical manifestations of HFpEF are atypical, and the pathogenic mechanism remains poorly understood, limiting clinical diagnosis and therapeutic options[4,5]. Accordingly, early and accurate diagnosis of HFpEF and determination of prognosis can contribute to implementing appropriate interventions to slow or halt disease progression and improve long-term consequences.

Despite the fact that patients with HFpEF have a left ventricular (LV) ejection fraction (LVEF) of greater than or equal to 50%, cardiac contractile performance may still be damaged, especially in individuals with severe concentric cardiac hypertrophy or a small LV cavity[6]. LVEF, a measure of overall cardiac systolic function derived from two-dimensional echocardiographic images, may be limited by altered cardiac geometry, boundary tracking difficulties, and other factors[7,8]. Compared with LVEF measurement, myocardial strain, which represents the percentage deformation of the myocardium in three directions (longitudinal, circumferential, and radial) during the cardiac cycle, is an emerging metric allowing more precise quantification of global and regional myocardial contractile function[9]. LV global longitudinal strain (LV GLS), which can be measured by speckle-tracking echocardiography (STE) or feature-tracking cardiovascular magnetic resonance (FT-CMR), is a commonly used myocardial strain parameter in cardiovascular diseases[10]. In recent years, multiple clinical trials have analyzed the diagnostic or prognostic significance of LV GLS in HFpEF, but they provided inconsistent results. In this situation, we implemented a meta-analysis to estimate the diagnostic and prognostic value of LV GLS in HFpEF.

## 2 Materials and methods

### 2.1 Literature Search Strategy

This systematic review and meta-analysis was performed depending on the principles of the Preferred Reporting Items for Systematic Reviews and Meta-Analysis 2020 statement (PRISMA)[11]. Two investigators (Shi and Wang) systematically performed documentary searches in four electronic databases (PubMed, Medline, Scopus, and Web of Science). English-language literature published from the inception of each database until December 20, 2022, was searched. Terms related to “Heart Failure, Diastolic”, “Heart Failure with Preserved Ejection Fraction”, “Heart Failure with Normal Ejection Fraction”, “Diastolic Dysfunction”, “Preserved Ejection Fraction”, “Global Longitudinal Strain”, and “GLS” were utilized following the rules of each database. For PubMed, the following search was performed: (((((Heart Failure, Diastolic[MeSH Terms]) OR (Heart Failure with Preserved Ejection Fraction[Title/Abstract])) OR (Heart Failure with Normal Ejection Fraction[Title/Abstract])) OR (Diastolic Dysfunction[Title/Abstract])) OR (Preserved Ejection Fraction[Title/Abstract])) AND ((Global Longitudinal Strain[Title/Abstract]) OR (GLS[Title/Abstract]))

### 2.2 Literature Inclusion and Exclusion Criteria

The inclusion criteria for this study were as follows: (i) diagnostic criteria: Fulfilment of diagnostic criteria for HF, patients with HFpEF had an LVEF≥45%, while HFrEF patients had an LVEF ≤40%[12,13]; (ii) study design: observational studies like case-control studies, prospective and retrospective cohort studies; (ii) endpoints: difference in LV GLS values between HFpEF patients and control groups (health controls or HFrEF patients), as well as the connection between LV GLS and diagnosis or adverse endpoints of HFpEF. The exclusion criteria for this study were as follows: (i) irrelevant or duplicated studies; (ii) the papers were case reports, reviews, letters, commentaries, editorials, or non-human studies; and (iii) the articles lacked full text or sufficient crude data.

### 2.3 Literature Quality Evaluation and Data Extraction

The quality estimation of the enrolled studies was evaluated by two independent reviewers (Xiong and Liu) using the Newcastle–Ottawa Quality Assessment Scale (NOS) system, a “star-based” grading system comprised of three parts (selection, comparability, and outcomes). A total NOS score ranged from 0 to 9, and studies achieving a score of 6 or above were considered high quality (Supplementary Table 1).

The required data from the included research was extracted and tabulated in specifically constructed Microsoft Excel spreadsheets for analysis. The extracted contents were as follows: (i) publication details: last name of the first author, year of publication, and the country setting; (ii) demographic characteristics: sample size, proportions of males, and mean age; (iii) study details: study design, LV GLS measurement method, data available to calculate the standardized mean difference (SMD) of LV GLS values between HFpEF patients and control groups (sample size, mean of LV GLS values and standard deviation), data related to diagnostic meta-analysis (LV GLS cut-off, area under the curve (AUC), sensitivity (SEN), specificity (SPE), true positive (TP), false positive (FP), false negative (FN), true negative (TN)), and data associated with prognostic meta-analysis (variables adjusted, follow-up duration, endpoints (HF hospitalization, cardiovascular (CV) hospitalization, CV death, and All-cause death), hazard ratios (HRs) and 95% confidence intervals (CIs)); and (iv) NOS quality scores. Two independent researchers (Yang and Qiao) conducted data extraction, and disagreements were resolved by mutual coordination or third-party adjudication (Dong and Liu).

### 2.4 Statistical Analysis

Meta-Disc (Version 14.0) was used to analyze the diagnostic value of LV GLS in HFpEF. The pooled SEN, SPE, positive likelihood ratio (PLR), negative likelihood ratio (NLR), diagnostic odds ratio (DOR), and AUC for the summary receiver operating characteristic curves (SROC) were used to characterize the diagnostic performance. Review Manager (Version 5.4) was used to analyze the difference in LV GLS values between HFpEF patients and control groups. SMD and 95% CIs expressed the pooled effect sizes. STATA (Version 16.0) was used to assess the association between LV GLS and adverse outcomes of HFpEF. HRs and 95% CIs represented the pooled effect sizes. The heterogeneity among the included studies was appraised by the Cochran Q statistics (chi-square test) and quantified with the I2 statistic. The fixed-effect model was employed when the Q test (I^2^<50%, p>0.05) revealed no significant heterogeneity across studies. When the Q test (I^2^≥50% or p<0.05) found prominent heterogeneity among studies, the random-effect model was utilized, followed by a Galbraith plot to explore the source of heterogeneity. Publication bias was evaluated using Deeks’ funnel plot, Funnel plot, and Egger’s test. Sensitivity analysis was employed to assess the impact of single research on the overall estimate by omitting one study each time. P<0.05 was considered to be statistically significant.

## 3 Results

### 3.1 Literature Search Results

A flowchart of the database search and text screening procedure is demonstrated in Fig 1. 2,492 publications were retrieved through database searching, consisting of 372 from PubMed, 624 from Medline, 559 from Scopus, and 937 from Web of Science. After excluding 874 duplicates, we screened the titles and abstracts of 1,618 papers. 1,524 articles were removed following the inclusion and exclusion criteria. Finally, among the remaining 94 studies, two independent researchers (Shi and Wang) read the full text and excluded 53 records due to repetitive research, irrelevant results, and insufficient data. Overall, 41 articles in total were enrolled in the meta-analysis.

**Fig.1.**
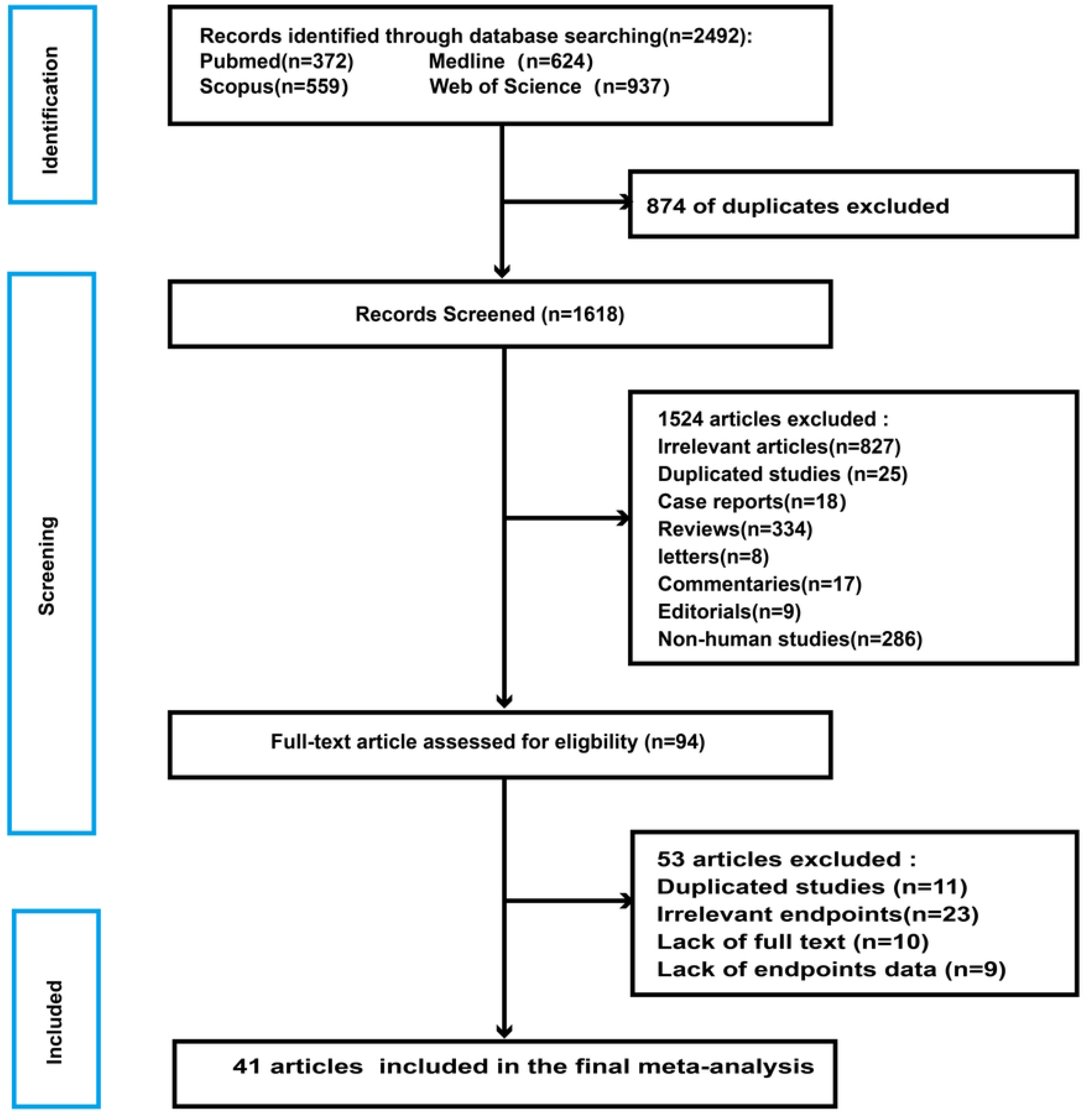
Flowchart of Database Search and Text Screening Procedure

### 3.2 Characteristics of Included Studies

Forty-one studies [14-53] published from 2008 to 2022 were investigated, including 27 case-control studies, 12 prospective cohort studies, and 2 retrospective cohort studies. Table 1 shows the baseline features of the selected research. A total of 14,543 patients (10,193 HFpEF patients, 6,931 males, mean age of 67.66 years) were involved. Thirty-two studies detected LV GLS using STE, and the other 9 utilizing FT-CMR. Thirty-eight studies reported mean LV GLS values in HFpEF patients (−8.2% - -20.8%), whereas the remaining 3 informed the median values (−15.3% - -20%). Concerning endpoint events, 27 studies evaluated the differences in LV GLS values between HFpEF patients and healthy controls (21 research) or HFrEF patients (16 research), and 26 studies assessed the diagnostic (6 trials) and prognostic (18 trials) significance of LV GLS in HFpEF. The NOS scores of included research varied from 6 to 9, indicating that the methodological quality was generally reliable (**S1 Appendix**).

**Table 1.**
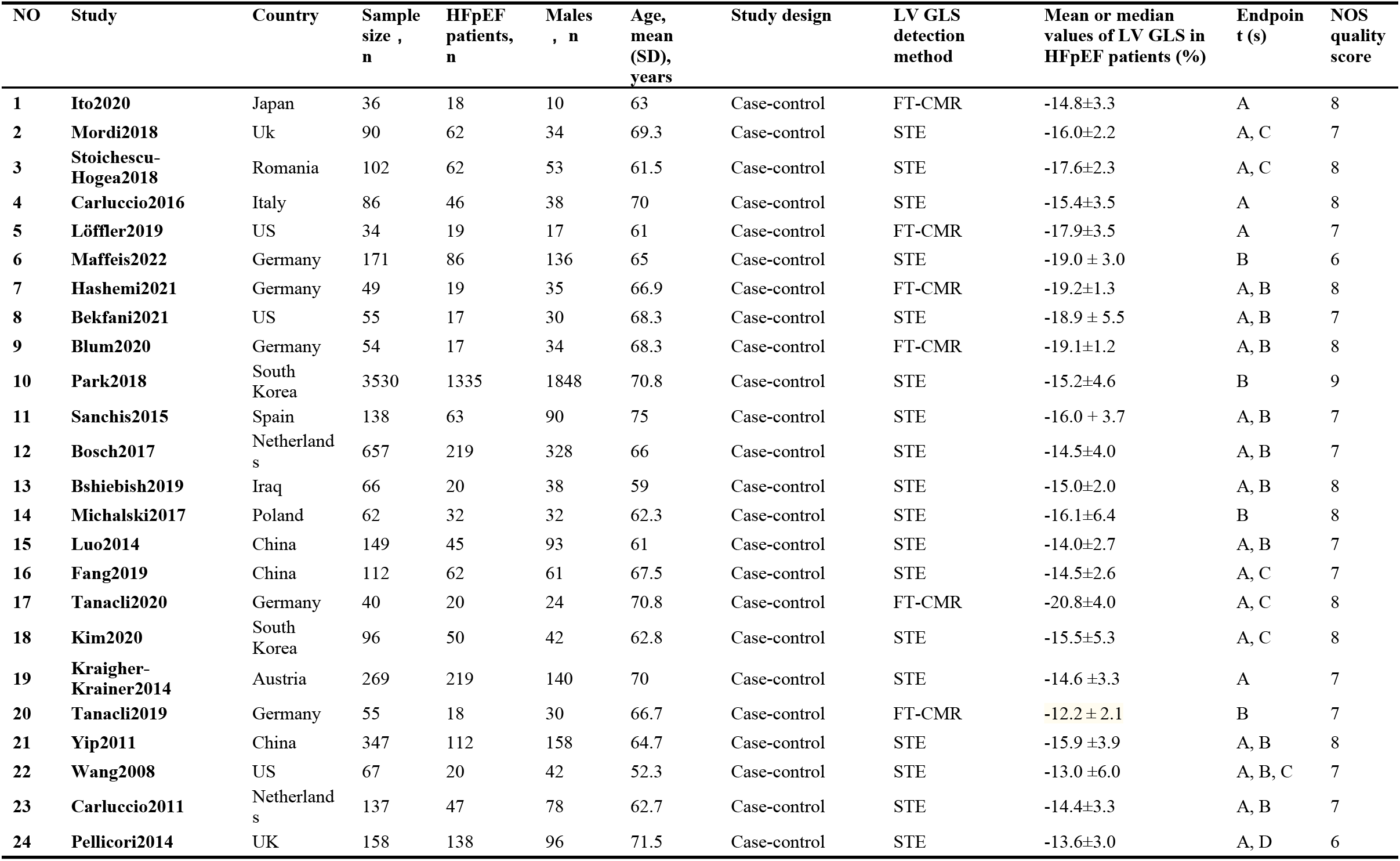
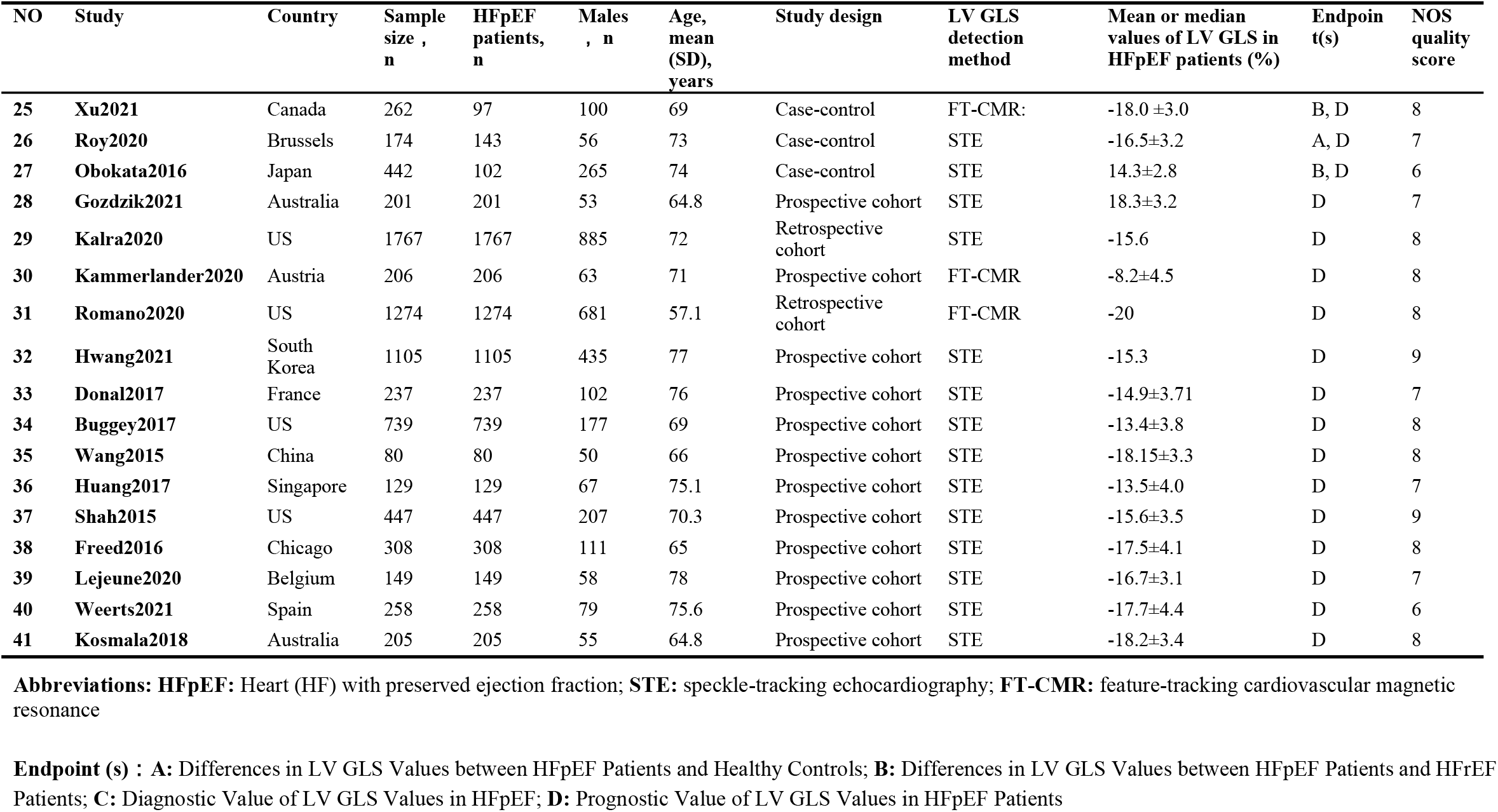
Baseline characteristics of the 41 selected research.

### 3.3 **Difference**s in LV GLS Values Between HFpEF Patients and Control Groups

Twenty-one studies[14–34] explored the differences in LV GLS values between HFpEF patients (n=1431) and healthy controls (n=864), with a mean LV GLS of - 16.07%±3.32% vs. -19.54%± 2.57% (Fig 2A). Moreover, 16 studies[14–18,21,26,30,33– 40] examined discrepancies in LV GLS values between HFpEF patients (n=2,234) and HFrEF patients (n=3,355), with an average LV GLS of -16.42%±3.56% vs. -9.3%±3.48% (Fig 2B). Meta-analysis using a random-effect model revealed that the LV GLS values in HFpEF patients were significantly lower than in healthy individuals (SMD:1.21; 95% CI (0.94, 1.47), p<0.00001, I^2^=85%; P<0.00001) (Fig 2C), but substantially higher than in HErEF patients (SMD: -2.03; 95% CI (−2.23, -1.72), p<0.00001, I^2^=92%; P<0.00001) (Fig 2D). According to the Galbraith radial plots, the research by (Fang et al., Wang et al., Sanchis et al.) and (Maffeis et al., Carluccio et al., and Yip et al.) were sources of heterogeneity for the above two comparison groups, respectively (S2 Appendix. 1A, 2A).

**Fig.2.**
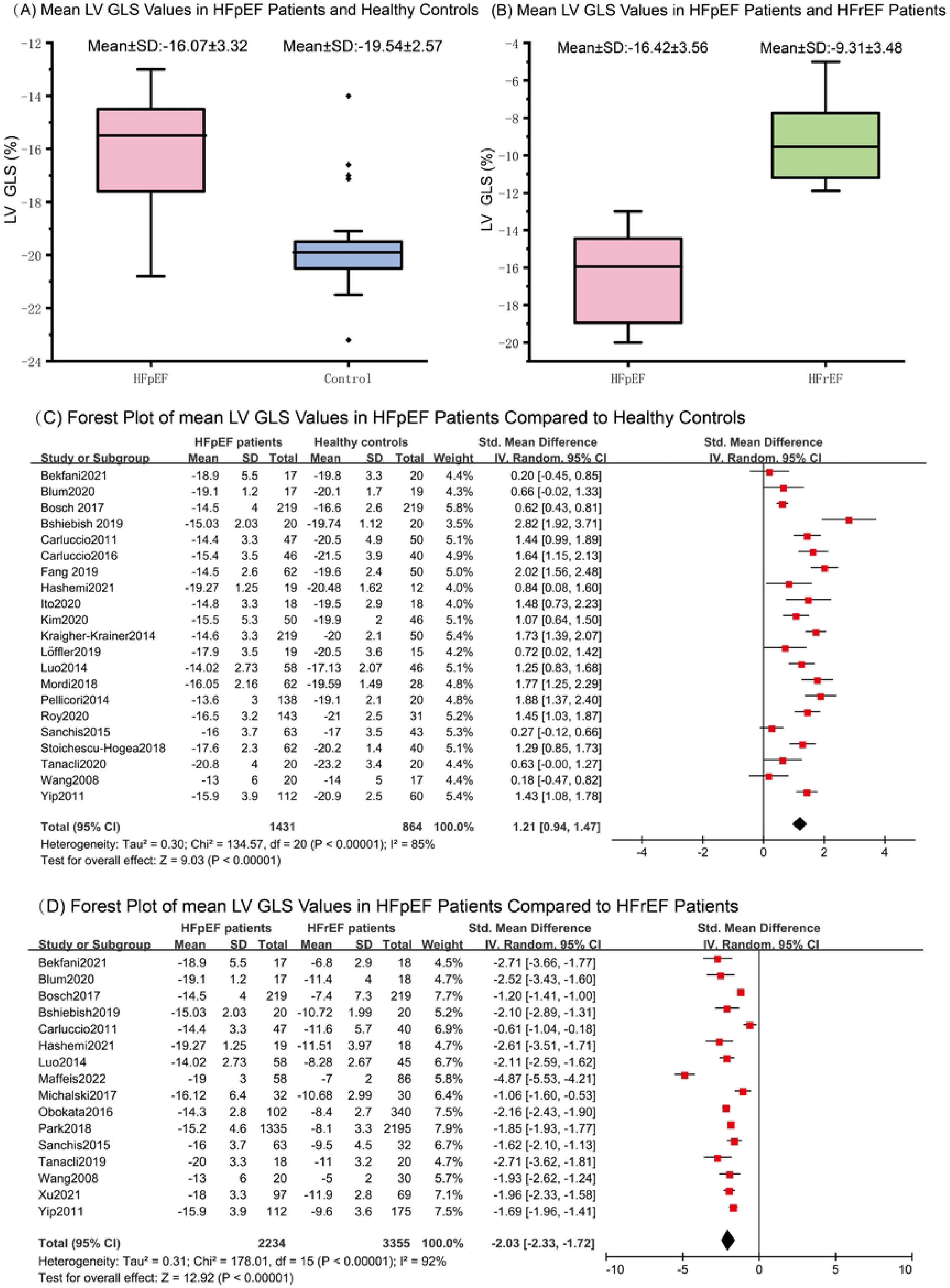
Differences in LV GLS Values between HFpEF Patients and Control Groups (A) Mean LV GLS Values in HFpEF Patients and Healthy Controls (B) Mean LV GLS Values in HFpEF Patients and HFrEF Patients (C) Forest Plot of mean LV GLS Values in HFpEF Patients Compared to Healthy Controls (D) Forest Plot of mean LV GLS Values in HFpEF Patients Compared to HFrEF Patients

The associated Funnel plots were asymmetrical (S2 Appendix. 1B, 2B) and the p-values of Egger’s test were 0.2 and 0.41 (S2 Appendix. 1C, 2C), indicating a potential publication bias among included studies. The sensitivity analyses removing one research each time indicated that none of the individual trials substantially impacted the pooled estimates (S2 Appendix. 1D, 2D). Overall, the results of this study were relatively constant.

### 3.4 Diagnostic Value of LVGLS in HFpEF

Six studies[20,23,27,31–33] estimated the diagnostic usefulness of LV GLS values in HFpEF compared with healthy controls, with LV GLS cut-off values ranging from -16% to -24.1 and AUC from 0.68 to 0.98 (Table 2). Five trails provided complete data for diagnostic meta-analysis (SEN, SPN, TP, FP, FN, and TN). The SROC curve revealed no typical “shoulder-arm” pattern, while the Spearman correlation coefficient between the logarithm of sensitivity and the logarithm of (1-specificity) was -0.1, P=0.873, suggesting no threshold effect in the meta-analysis. Summary assessments of the diagnostic performance of LV GLS in HFpEF were as follows: SEN was 0.77 (95% CI: 0.71–0.82) (Fig 3A); SPE was 0.66 (95% CI: 0.58–0.74) (Fig 3B); PLR was 2.17 (95% CI: 1.36– 3.47) (Fig 3C); NLR was 0.35 (95% CI: 0.21–0.59) (Fig 3D); DOR was 7.53 (95% CI: 3.19–17.74) (Fig 3E); AUC for the SROC was 0.81 (95% CI: 0.79–0.87) (Fig 3F). The p-value of Deeks’ test (0.18) was greater than 0.05, suggesting no significant publication bias. (S2 Appendix. 3).

**Table 2.** Data related to diagnostic meta-analysis.

|  | **Study** | **LVGLS cut-off (%)** | **AUC** | **SEN (%)** | **SPE(%)** | **TP** | **FP** | **FN** | **TN** | **P-value** |
| --- | --- | --- | --- | --- | --- | --- | --- | --- | --- | --- |
| 1 | Mordi2018 | -17.8 | 0.78 | 0.83 | 0.63 | 52 | 10 | 11 | 17 | 0.037 |
| 2 | Stoichescu-Hogea2018 | -19.35 | 0.83 | 0.79 | 0.47 | 49 | 21 | 13 | 19 | <0.001 |
| 3 | Fang2019 | — | 0.87 | — | — | — | — | — | — | <0.001 |
| 4 | Tanacli2020 | -24.1 | 0.69 | 0.90 | 0.41 | 18 | 12 | 2 | 8 | 0.037 |
| 5 | Kim2020 | -16.7 | 0.68 | 0.54 | 0.85 | 27 | 7 | 23 | 39 | 0.001 |
| 6 | Wang2008 | -16 | 0.98 | 0.95 | 0.95 | 19 | 1 | 1 | 16 | <0.001 |

**Abbreviations:** LV GLS: left ventricular global longitudinal strain **TP:** True positive; **FP:** False positive; **FN:** False negative; **TN:** True negative; **SEN:** Sensitivity; **SPE:** Specificity.

**Fig.3.**
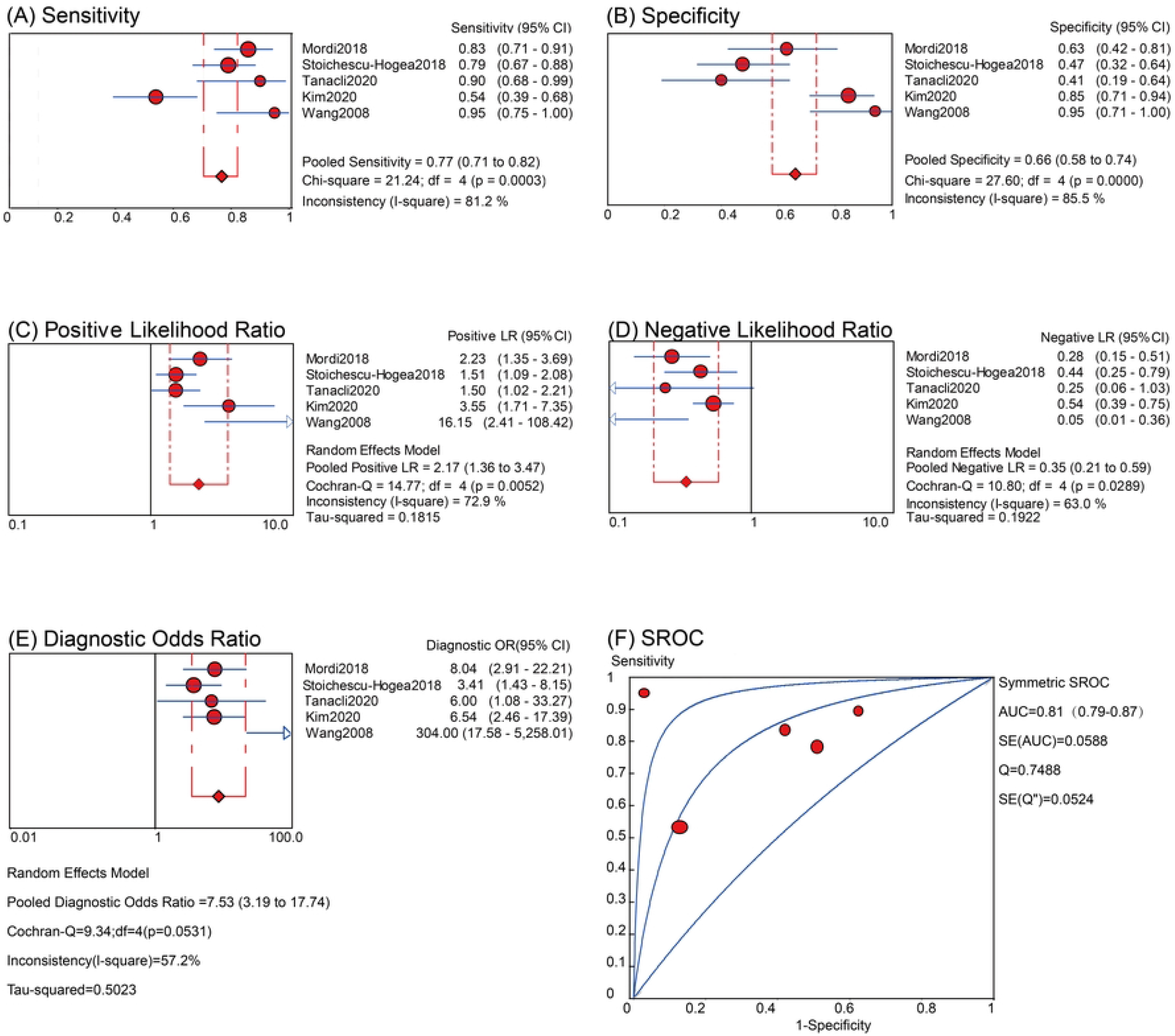
Diagnostic Value of LVGLS in Patients with HfpEF (A) Sensitivity (B) Specificity (C) Positive Likelihood Ratio (D) Negative Likelihood Ratio (E) Diagnostic Odds Ratio (F) SROC

### 3.5 Prognostic Value of LV GLS in HFpEF Patients

Eighteen studies[6,28,37,40–53] examined the connection between LV GLS values as a continuous variable and adverse outcomes in HFpEF patients. The duration of follow-up varied from 4 to 144 months. In 9 studies, variables such as age, sex, race, history of CV disease, medication history, laboratory tests, and others that might affect the association between LV GLS and research outcomes were adjusted (Table 3). Meta-analyzes with random-effects models showed that the low LV GLS values in HFpEF patients were associated with a higher risk of all-cause death (HR: 1.12; 95% CI: 1.01-1.25; p=0.000, I^2^=84%; P = 0.031)[29,42,44,47,49,51] (Fig 4A), but not with the composite outcomes (all-cause death and HF hospitalization (HR: 1.03; 95% CI: 0.95-1.11; p=0.000, I^2^= 83%; P = 0.51)[29,45–48,51,52] (Fig 4B) or CV death and HF hospitalization (HR: 1.04; 95% CI: 0.94-1.14; p=0.000, I^2^= 81%; P = 0.49)) [6,28,37,41,43] (Fig 4C)). Additionally, three studies by Xu et al., Freed et al., and Kosmala et al. reported no association between low LV GLS values in HFpEF patients and the composite outcome of all-cause death and CV hospitalization [40,50,53]. The result for all-cause death showed substantial heterogeneity. The Galbraith radial plot suggested that the studies by Romano2020 et al. and Buggey2017 et al. may be the source of heterogeneity (S2 Appendix. 4A). the associated Funnel plot was asymmetric (S2 Appendix. 4B) and the p-value of Egger’s test was 0.67 (S2 Appendix. 4C), suggesting a possible publication bias across included studies. The sensitivity analysis indicated that none of the studies significantly affected the pooled estimates (S2 Appendix. 4D).

**Table 3.**
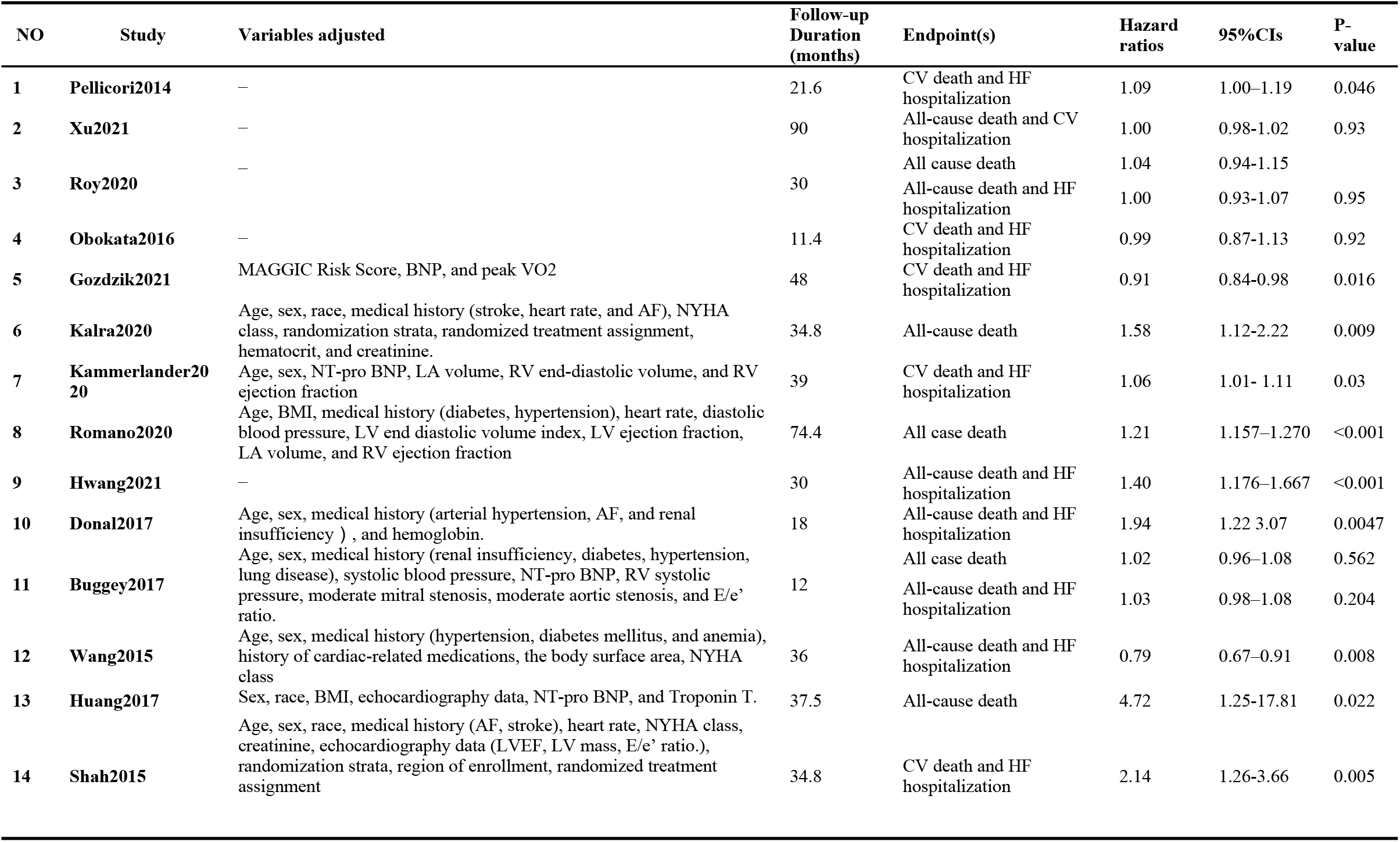
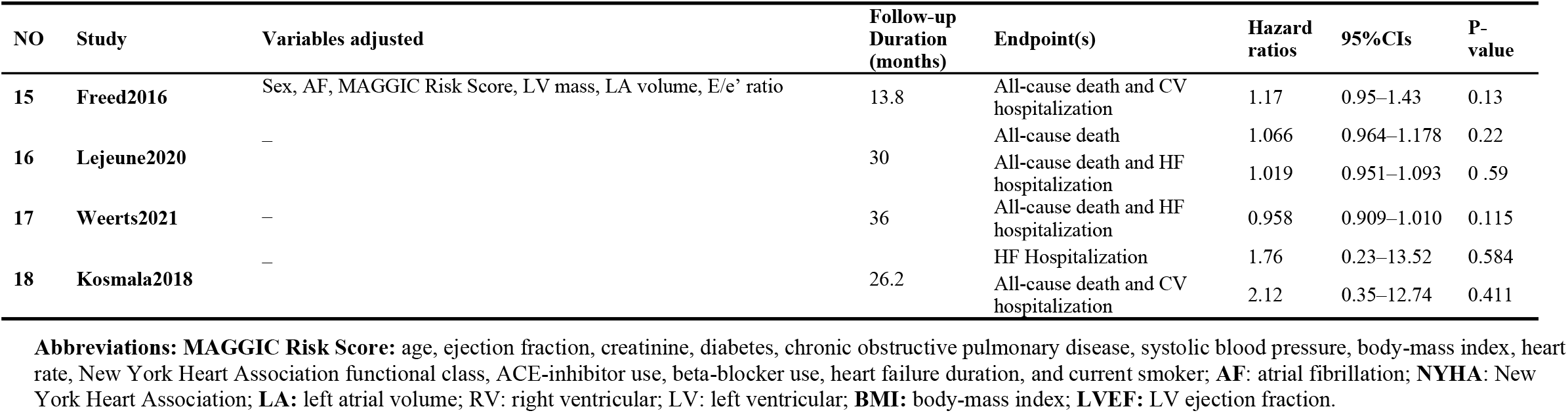
Data related to Meta-analysis of prognostic assessment.

**Fig.4.**
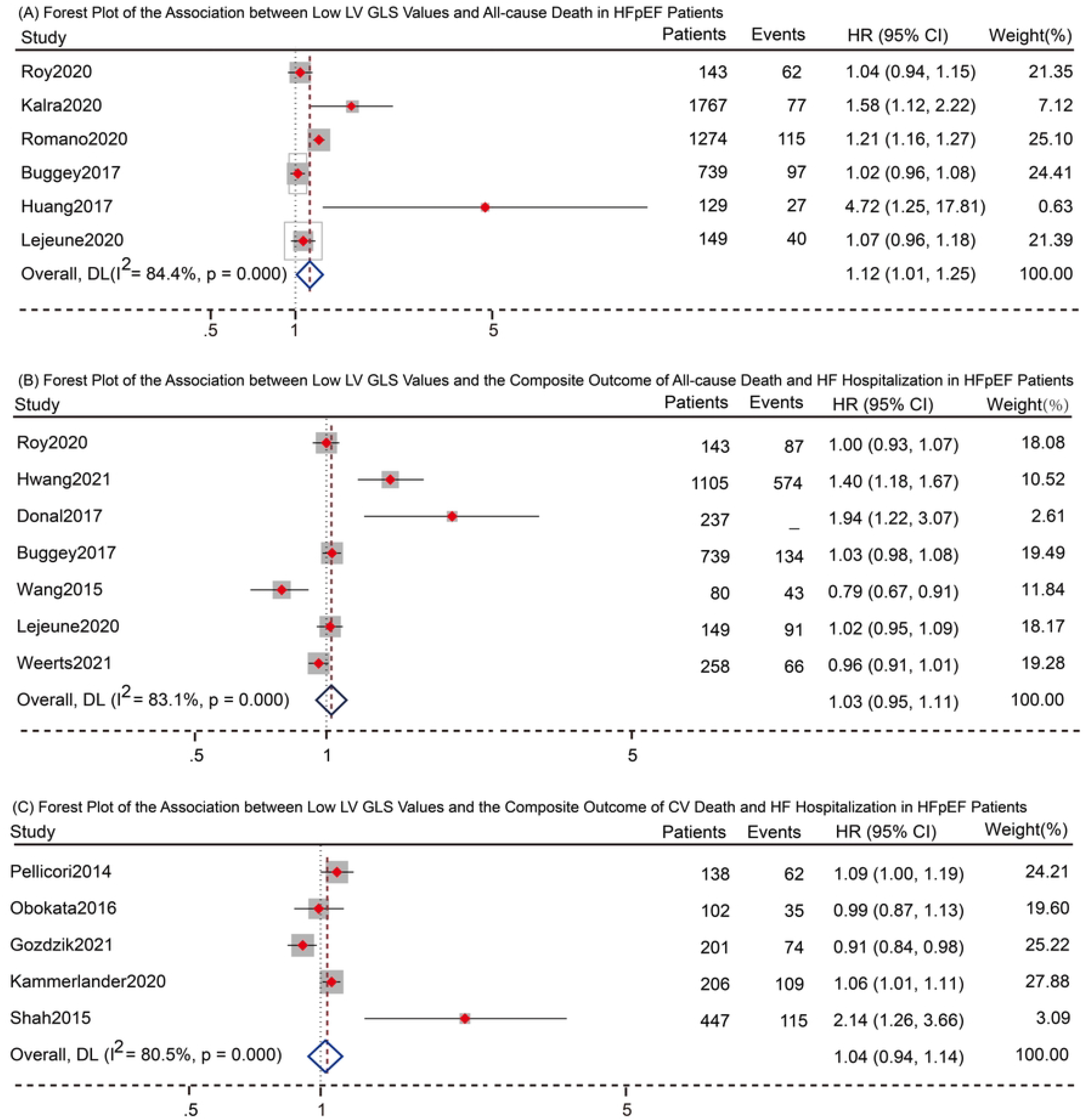
Associations between Low LV GLS Values and Adverse Outcomes in HFpEF Patients (A) Forest Plot of the Association between Low LV GLS Values and All-cause Death in HFpEF Patients (B) Forest Plot of the Association between Low LV GLS Values and the Composite Outcome of All-cause Death and HF Hospitalization in HFpEF Patients (C) Forest Plot of the Association between Low LV GLS Values and the Composite Outcome of CV Death and HF Hospitalization in HFpEF Patients

## 4 Discussion

Current European Society of Cardiology guidelines graded HF based on LVEF into reduced (LVEF≤40%), mildly reduced (LVEF 41-49%), and preserved (LVEF≥50%) three subtypes[13]. HFpEF, which accounts for about half of all HF, is evolving as an increasingly severe public health concern[2]. Presently, HFpEF is diagnosed depending on representative symptoms and/or signs of HF (breathlessness, fatigue, edema, and others), an LVEF of more than 50%, objective evidence of cardiac structural or functional abnormalities, and a growing level of BNP or NT pro-BNP[5]. However, most sufferers lack specific clinical manifestations in the initial stages and are usually not detected until the impairment of cardiac structure and function develops [54]. It is essential to explore effective metrics for identifying high-risk individuals, early diagnosis, prognosis assessment, and treatment monitoring.

During cardiac systole, the obliquely and oppositely oriented subendocardial and epicardial myofibers produce apical counter-clockwise torsion and basal clockwise torsion, causing the LV to shorten in the longitudinal and circumferential planes while thickening in the radial plane[55]. Cardiac strain assesses the change in length of the myocardium in three planes (longitudinal and circumferential (expressed by a negative value), radial (represented by a positive value) relative to the initial length during the cardiac cycle. It can be calculated using the following formula: strain (epsilon) = L-Lo/Lo (where Lo is the baseline length of the myocardium, and L is the length after deformation)[7]. LV GLS, a sensitive and objective diagnostic indicator in cardiac strain imaging, can detect LV mild contractile dysfunction before changes in LVEF[8]. It is currently used as an auxiliary parameter for diagnosis and prognostic assessment in cardio-oncology[56] and is being expanded to other fields such as cardiac amyloidosis[57], cardiomyopathies[58], valvular heart diseases[59], pulmonary hypertension[60], and HF[58]. Although numerous studies have confirmed the presence of abnormal LV GLS in HFpEF patients and the Heart Failure Association consensus has unanimously recommended impaired V GLS as an ancillary criterion for the diagnosis of HFpEF[54], there is controversy about whether LV GLS has independent diagnostic and prognostic value for HFpEF. We consequently performed this meta-analysis to assess the diagnostic and prognostic value of LV GLS in HFpEF.

The results of the current meta-analysis illustrated a significant link between LV GLS and the diagnosis and prognosis of HFpEF. First, the LV GLS values in HFpEF patients were significantly lower than in healthy individuals, but substantially higher than in HErEF patients, confirming the presence of mild LV systolic dysfunction in HFpEF patients. The longitudinal systolic function of LV is determined by the endocardium, which is highly susceptible to the detrimental impacts of ischemia or hypertrophy[61,62]. Multiple comorbidities in HFpEF patients may promote microvascular malfunction and muscle fibrosis[63], reducing LV longitudinal systolic function and ultimately leading to LV GLS impairment. Second, LV GLS has excellent auxiliary diagnostic value for HFpEF, with with an AUC of 0.81 (95% CI: 0.79–0.87). Finally, the low LV GLS values in HFpEF patients were correlated with a higher risk of all-cause death, indicating that LV GLS is a powerful independent predictor of all-cause mortality in HFpEF patients.

This meta-analysis involved a relatively comprehensive literature search, and the included studies were of excellent quality according to the NOS score. Nonetheless, some limitations must be considered: First, the heterogeneity of the included studies was high, with probable causes including the detection method of LV GLS, the diagnostic criteria of HF, and the study design. Second, in the diagnostic meta-analysis, the diagnostic cut-off values of LV GLS were not consistent, which might be related to factors like the detection technique. Third, in the prognostic meta-analysis, the follow-up periods were inconsistent, and the number of studies in which effect sizes could be combined for the same endpoint was low, limiting the ability to draw significant conclusions. As a result, our meta-analysis is exploratory, and further high-quality original research is needed to support the findings of this study.

## 5 Conclusion

LV GLS is impaired in HFpEF patients despite a normal left ventricular ejection fraction, indicating the existence of mild LV contractile dysfunction. Moreover, LV GLS might be an auxiliary indicator for diagnosing HFpEF and predicting all-cause death in HFpEF patients.

## Data Availability

All relevant data are within the paper.

## 6 Supporting information

**S1 Appendix. NOS score of the included studies (PDF)**

**S2 Appendix. Heterogeneity, publication bias, and sensitivity analysis of relevant meta-analyses (PDF)**

**S3 Appendix.PRISMA checklist (PDF)**

## 7 Acknowledgments

The authors would like to express their gratitude to Professor W. Szymański for his valuable help in the process of retrieving relevant literature.

## 8 Author Contributions

**Conceptualization:** Yujiao Shi, Guoju Dong, Jiangang Liu

**Data curation**: Yujiao Shi, Wang Qing

**Formal analysis:** Wang Qing, Chunqiu Liu, Xiong Shuang

**Investigation:** Chenguang Yang, Wenbo Qiao

**Methodology:** Yujiao Shi, Guoju Dong, Jiangang Liu

**Project administration:** Wang Qing, Chunqiu Liu, Xiong Shuang

**Supervision:** Guoju Dong, Jiangang Liu

**Visualization:** Yujiao Shi, Chunqiu Liu

**Writing – original draft:** Guoju Dong, Jiangang Liu

**Writing – review & editing:** Yujiao Shi, Guoju Dong, Jiangang Liu

## 9 Data availability statement

All relevant data are within the paper.

